# Superposition of Droplet and Aerosol risk in the transmission of SARS-CoV-2

**DOI:** 10.1101/2022.09.28.22280473

**Authors:** John E. McCarthy, Barry D. Dewitt, Bob A. Dumas, James S. Bennett

## Abstract

**Objectives:** Considering three viral transmission routes: fomite contact, aerial transmission by droplets, and aerial transmission by aerosols, the aerial routes have been the focus of debate about the relative role of droplets and aerosols in SARS-CoV-2 infection. We seek to quantify infection risk in an enclosed space via short-range airborne transmission from droplets and long-range risk from aerosols toward focusing public health measures.

**Methods:** Data from three published studies were analyzed to predict relative exposure at distances of 1 m and farther, mediated by droplet size divided into two bins: larger than 8 *µm* and smaller than 75 *µm* (medium droplets) and smaller than 8 *µm* (small droplets or aerosols). The results at 1 m from an infectious individual were treated as a boundary condition to model infection risk at greater distance. At all distances, infection risk was treated as the sum of exposure to small and medium droplets. It was assumed that number of virions is proportional to droplet volume.

**Results:** The largest infection risk (as exposure to droplet volume) came from medium droplets, close to the infectious individual out to approximately 1 m. Farther away, the largest risk was due to aerosols. For one model, medium droplet exposure disappeared at 1.8 m.

**Conclusions:** Policy concerning social distancing for meaningful infection reduction relies on droplet exposure as a function of distance, yet within this construct droplet size determines respiratory deposition. This two-fold distance effect can be used to evaluate additional measures such as plexiglass barriers and masking.

## 1 Introduction

There are believed to be three transmission routes for severe acute respiratory syndrome-coronavirus 2 (SARS-CoV-2): touching eyes, nose, or mouth with a hand that has touched a fomite (infected surface or person), aerial transmission by droplets, and aerial transmission by aerosols [3]. The last of these has been the subject of debate but is likely important[7], including the distinction between “droplets” and “aerosols” and their relative risk of causing infection [12, 2].

Although it is now clearer to the broader research community and the public that the threshold between “droplet” and “aerosol” is dynamic [12], depending on both the pathogen and environmental conditions, the relative importance of droplets and aerosols to infection risks, and the consequences for mitigation policies, has not been well studied. A report from the UK’s Scientific Advisory Group for Emergencies estimated the risk of infection for a non-infected person standing at 1m from an infectious person to be at most an order of magnitude larger than the risk of infection at double that distance [13]. We endeavor to improve on estimates of that kind, by comparing the relative viral loads received through standing close to an infectious person with the amount received more generally indoors, such as in classrooms, airplanes, and stadiums. We leverage results from previous empirical research to better model infection risk, potentially as input to risk-cost-benefit analyses of common activities for public and private decision-making [10].

A historical dismissal of air-quality related strategies to mitigate airborne pathogens in general [11], means that many built environments can be loci of SARS-COV-2 spread. Here, we attempt to estimate the risks of aerial transmission, operationalized as the quantity of virus in particles exhaled by an infected person and inhaled by a currently uninfected person. We first describe our terminology and the conceptual model. We then incorporate data from the literature to estimate viral loads due to aerial transmission and include estimates derived from bacteriophage droplet tracer studies, for possible exposure during a commercial flight. Finally, we discuss implications for policymaking.

## 2 Methods

### 2.1 Droplets and Virions

When an infected person exhales (or vocalizes, coughs or sneezes) they spray *droplets* that contain the virus into the air. The medium droplets collide with a person, land on a surface, or fall to the ground. The small droplets or aerosols can waft through the air for hours, travel a long distance, and potentially reach the respiratory tract of a distant person. We estimate the number of virions dispersed via the respiratory tract by taking this number to be proportional to the initial droplet size as volume. Then, we compare those quantities at different dis-tances, to get a better estimate of the exposure, by making a chain of inferences, explained at more length below.

### 2.2 Data from Literature

The terms “droplet” and “aerosol” are used in the literature with respect to whether they pose *short-range* or *long-range* aerial transmission risk. It is desirable to not require a constant size threshold to separate the particles defined by the two terms, the latter being a common, but much-criticized, practice [12]. In this study, we do apply a size threshold to align with cited literature that separates droplets into small and medium bins.

In Subsection 2.5 we use experimental results of Shah et al. [14] to estimate the long-range risk from aerosols for a given rate of aerosol emission. In Subsection 2.4 we use theoretical results of Chen et al. [5] to estimate the short-range risk from emitted droplets. We then use experimental results of Duguid [6] and Chao et al. [4] to estimate what fraction of exhaled particles is aerosolized (i.e., contributes to long-range risk) and what fraction contributes to short-range risk (as droplets). Subsection 3.1 compares the risk at distances 1m versus 2m.

### 2.3 Alice and Bob

To provide definitions, list assumptions, and outline the analytical procedure, we use the following scenario. An infectious individual, whom we shall call Alice, poses an airborne infection risk to a non-infected individual, whom we shall call Bob.

1. Bob is in an enclosed environment with Alice for an extended period of time (an hour or more). Alice is exhaling saliva and lung fluid droplets of different sizes, all with the same expected concentration of virions per unit volume, initially, at launch. We classify Alice’s droplets into two size bins.
2. Droplets with diameter between 8 and 75 *µm* we call *medium droplets*. These are pulled down by gravity and pose risk to Bob if they land in his mouth, nose or eyes. We call this Route 1.
3. Droplets with diameter between 1 and 8 *µm* we call *aerosols or small droplets*. These drift in the air. The risk they pose to Bob is if he inhales them. We call this Route 2.
4. In Table 2, we give measurements from [5] and [6] of the ratio of volumes for total exhalation in Medium droplets to total exhalation in Small droplets. Let us call this ratio *ρ*. The fraction of the total emission of all droplets that is Small droplets is then 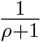
5. From [5], we get that every 1 *µL* of exhaled droplets produces 17×10^*−*6^*µL* of Medium droplets entering the respiratory tract via Route 1 if Bob is facing Alice at a distance of 1 *m*. We attribute all Route 1 exposure to these non-aerosolized droplets.
6. To extrapolate what the Route 1 droplet inhalation would be at different distances, we use two different models: a rapid decay model from [5] and an inverse distance square decrease.
7. For the Route 2 exposure, we use the experimental data of [14]. That study measures what fraction of a given aerosol emission is inhaled at a distance of 2*m*. We multiply this fraction by 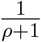to estimate the total inhalation of aerosols by Bob via Route 2 at a distance of 2 *m* for every 1 *µL* of droplets exhaled by Alice.
8. To extrapolate the aerosol inhalation (Route 2) at different distances, we use the bacteriophage measurements in aircraft cabins of [9].
9. We graph the medium droplet (blue, orange) and aerosol (gray, yellow) results, under different scenarios in Figures 1 and 2. To extrapolate the Route 1 exposure at various distances, we applied the Chen (blue) or inverse square (orange) functions to the estimate at 1*m*.
10. The total exposure of Bob is the sum of the Route 1 and Route 2 exposures. We assume that Bob’s risk of infection is proportional to the total exposure.

**Table 1:** Adapted from [14]. First column indicates whether the exhaling mannikin is wearing a mask. Second column is the number of Air Changes per Hour. Third column is the percentage of exhaled particles that arrive at the detector every hour. Fourth column is the rate at which particle concentration drops. Fifth column is the steady state saturation as a percentage of the emission rate.

| Mask | ACH | $R^*$ (% $h^{-1}$) | $\lambda^*$ ($h^{-1}$) | $c_{\text{sat}}^* = R^*/\lambda^*$ |
| --- | --- | --- | --- | --- |
| None | 0 | $0.53 \pm 0.11$ | $0.46 \pm 0.11$ | $1.13 \pm 0.057$ |
| Surgical | 0 | $0.41 \pm 0.36$ | $0.41 \pm 0.39$ | $0.99 \pm 0.11$ |
| None | 1.7 | 0.48 | 1.36 | 0.35 |
| None | 3.2 | 0.41 | 2.27 | 0.18 |

**Table 2:** Volume of various droplet sizes from [4] (C) and [6] (D).

|  | C Speak | C Cough | D Speak | D Cough | D Sneeze |
| --- | --- | --- | --- | --- | --- |
| Vol. in Small droplets | 3E-06 | 6E-06 | 6E-06 | 1E-04 | 4E-02 |
| Vol. in Medium droplets | 8E-04 | 1E-03 | 5E-03 | 9E-02 | 9 |
| Ratio (Medium/Small) | 270 | 160 | 760 | 786 | 200 |

**Figure 1:**
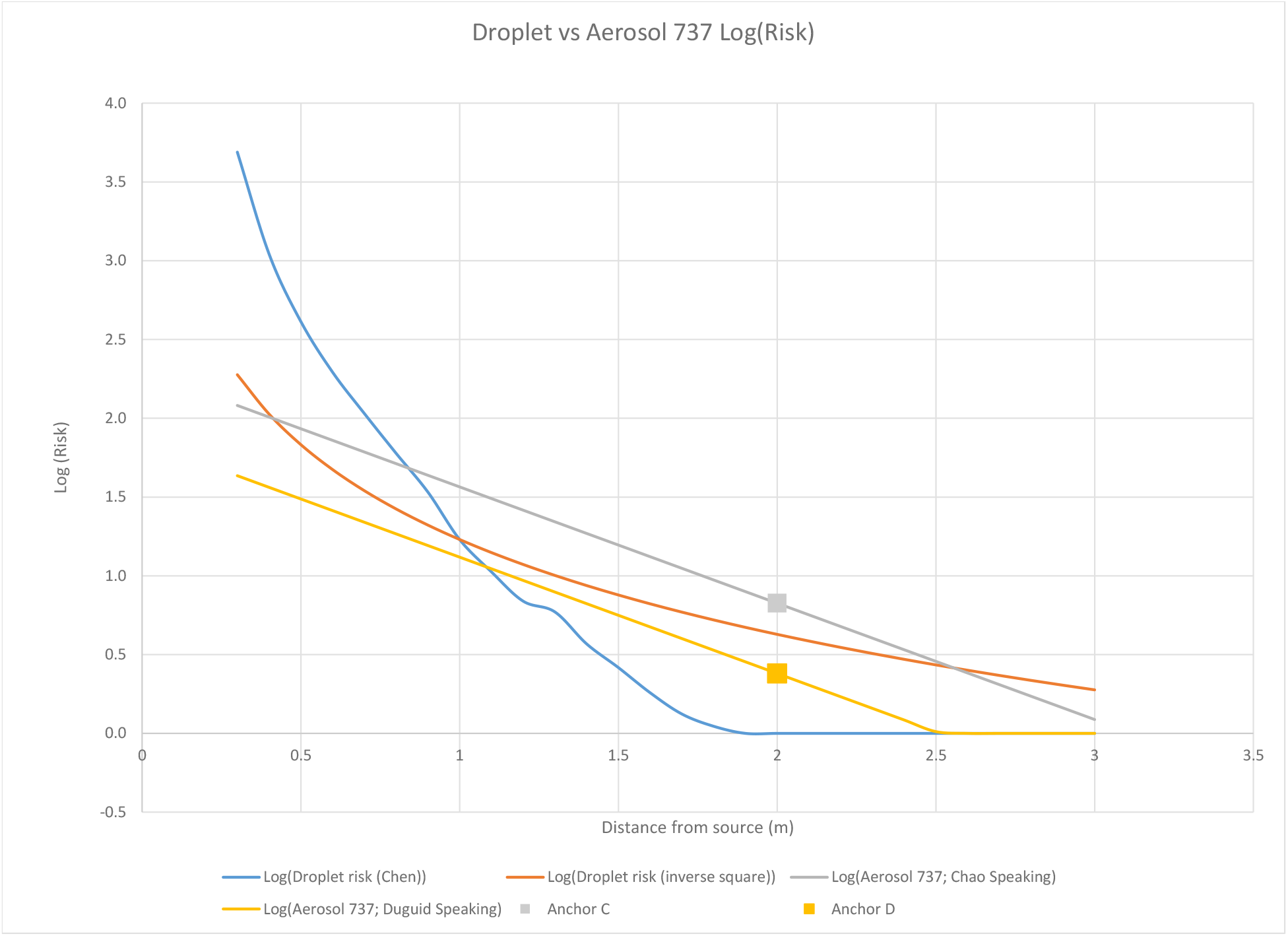
Decrease of risk, as virion exposure, with distance from an infectious person. The decrease measured in 737 mock-up tracer experiments is normalized or “anchored” to intersect the speaking data at 2m from Chao (gray) and Duguid (yellow). The anchor values from Table 3 are plotted on the log base 10 scale.

**Figure 2:**
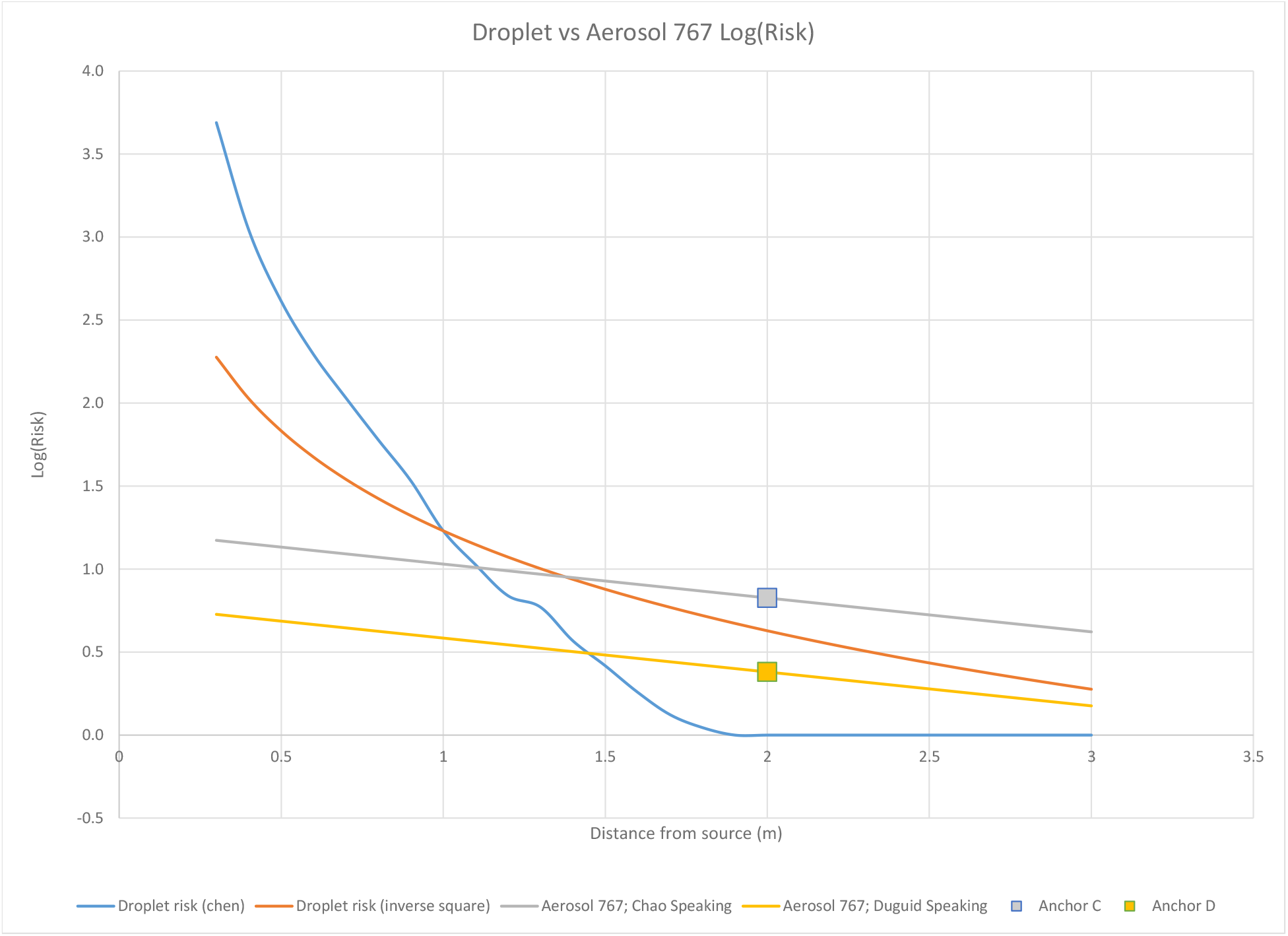
Decrease of risk, as virion exposure, with distance from an infectious person. The decrease measured in 767 mock-up tracer experiments is normalized or “anchored” to intersect the speaking data at 2m from Chao (gray) and Duguid (yellow). The anchor values from Table 3 are plotted on the log base 10 scale.

### 2.4 Droplet exposure—Route 1

In [5], Chen et al. analyze short-range droplet transmission, based on a mathematical model. They argue that there are two main routes of shortrange non-fomite transmission: large droplets that are projected directly into the mouth, nose and eyes of a nearby facing person at the same height (they ignore droplets that hit any other part of the face or body), and small droplets that enter the air stream and are inhaled. Large droplets are intrinsically short-range, because gravity pulls them down to the floor in a short period of time. (However, if coughed out, they can travel a long distance horizontally). They conclude that mid-size droplets (defined as having initial diameters 75-400 *µ*m) fall to the ground within a meter. Smaller droplets follow the airflow; larger ones decelerate horizontally more slowly, so travel farther, but will settle to the ground. Moreover, they conclude that at distances over 0.3m (talking) and over 0.8m (coughing) the majority of exposure comes from inhaled droplets rather than deposited droplets.

For their base data, [5] use a paper by Duguid [6] that measured the number and size of droplets exhaled by a person coughing, and by counting loudly from 1 to 100. The latter produced a total measured volume of 0.36 *µ*L (of which 2 *×*10^*−*3^ *µ*L came from droplets with a diameter less than 75*µ*m). The conclusion of [5] that we will use with respect to short-range aerial transmission is that <u>face-to-face, at a range of 1m, a person inhales 6.2 *×*10^*−*6^ *µ*L of the original 0.36 *µ*L of the talking emission, almost all of it from droplets smaller than 75 *µ*m.</u> Dividing by 0.36 we get that every 1 *µL*of exhaled droplets produces 17*pL* of inhaled droplets, via Route 1, from a facing subject at 1*m* (we change from *µL* to *pL* to make the numbers easier to read and compare).

We shall then multiply the number 17*pL* by a function depending on distance from the source to get the exposure at different distances. We shall refer to this technique of estimating the exposure at a specific distance and then multiplying by a function that decreases with distance as <u>anchoring</u>. The Route 1 distance functions decay more rapidly than those of Route 2.

### 2.5 Aerosol exposure—Route 2

In [14], Shah et al. set up a mannikin with a mechanical ventilator that exhaled atomized olive oil droplets, with a mean diameter of 1 *µ*m. The concentration *c* of oil in the air was measured for many hours at a distance of 2 m. Olive oil was chosen because its use with the experimental setup produced aerial particles of similar sizes to those produced during human exhalation. The study’s results fit well with the following mathematical model:

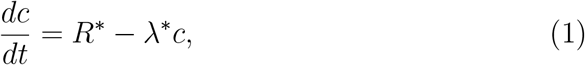

where *R*^*∗*^, which depends on where the concentration is measured, is a percentage of the particle injection rate *R*, and *λ*^*∗*^ is the particle decay rate, which is also dependent on location.

Equation (1) replaces the transport-diffusion equation with an assumption of instantaneous distribution of the aerosols. Its solution is given by

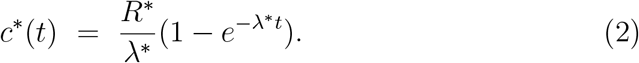

While the particle source is active, the quantity *c*^*∗*^(*t*) from Equation (2) will tend asymptotically to 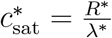

Shah et al. measured the output both with and without masks on the mannikin, and they also measured the output given different air change rates per hour (ACH). Table 1 summarizes some of their results for several masking and ACH combinations.

Thus, for example, at a distance of 2m, they found that the steady-state concentration was 1.13% (*±*.057 %) of the breath particle injection rate (final column, second row of Table 1). Note that even though the surgical masks were estimated to be 47% effective at blocking particles flowing through them, a significant amount of escaped around the bridge of the nose, thus diminishing their overall effectiveness.

Using the values of *R*^*∗*^ and *λ*^*∗*^ from Table 1 and equation 2, we get the estimate at 2m of

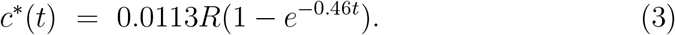

Equation 3 has little directional dependence: Shah et al. did measure at a distance of 2m and at angles of 0^*o*^, 90^*o*^ and 180^*o*^ from the source, and found the variation to be less than 10%.

## 3 Results

### 3.1 Comparing Route 1 and Route 2 exposures

Table 2 shows the aerosol exposure at 2m for a given emission rate.

From [5], we get that every 1 *µL* of exhaled droplets produces 17*pL* of inhaled droplets from a facing subject at 1*m*. We estimate the long-range risk by first using the numbers from Table 2 to estimate what fraction of that 1 *µL* is aerosolized, and then use the data from Table 1 to estimate how much of that is inhaled at steady state conditions at a distance of 2m. We assume an exhaled particle is aerosolized when it has a diameter under 8 *µm*, although environmental conditions change the diameter at which particles remain airborne [12].

These assumptions yields the following table. The columns use the measurements from [4] and [6]. The rows are the four conditions in Table 1.

The first entry, for example, is taken by dividing 1*µL* by 271, which Table 2 tells us is the fraction of the original emission that is aerosolized using the measurements from [4] and the assumption that only the small droplets become aerosolized, and then multiplying this number by 1.13% which comes from the last column in table 1.

### 3.2 Decay with distance

As far as we are aware, no one is certain how the risk from either droplets or aerosols decays with distance from the source. For droplets, we shall first use the theoretical model of [5]. This has a very rapid decay with distance.

To compare it with a more conservative estimate, we also model the decay is inverse square with distance. We shall anchor the latter with the same exposure at 1m from the Chen et al. model.

For aerosols, we shall use the study [9] by Lynch et al. that measured decay of aerosols with distance in planes. Their best fit for a single-aisle Boeing 737 was *e*^*−*1.7*x*^, where *x* is the distance from the source in meters, and for a dual-aisle Boeing 767 it was *e*^*−*.47*x*^. We shall anchor with the measurement from Table 3 with the largest measured ventilation, 3.2 ACH. The aircraft cabin exponential decay curves plotted on the log scale are the straight lines in Figures 1 and 2. In the absence of ventilation, the decay of aerosol risk with distance is likely to be much smaller—indeed, in small enclosed environments, the steady state may be fairly homogeneous.

**Table 3:** Steady-state aerosol intake in *pL* for every *µL* emitted, at a 2m distance from the source.

| Mask | ACH | Chao Speak | Chao Cough | Duguid Speak | Duguid Cough | Duguid Sneeze |
| --- | --- | --- | --- | --- | --- | --- |
| No | 0 | 41 | 68 | 14 | 14 | 55 |
| Surgical | 0 | 37 | 61 | 13 | 13 | 49 |
| No | 1.7 | 13 | 22 | 4.6 | 4.5 | 17 |
| No | 3.2 | 6.7 | 11 | 2.4 | 2.3 | 9 |

## 4 Discussion

Figures 1 and 2 show that effective interventions to reduce exposure vary dramatically for the short and long range routes. Even these rough estimates show the relative magnitudes of Medium droplet and Aerosol exposures as a function of distance. For example, Route 1 accounting for much of the risk close to a source indicates interventions such as plexiglass barriers between a customer and a store cashier would be important; in contrast, Route 2 dominating farther away shows how respirators, air filtration, and air disinfection would be paramount. Of course, interventions such as masks can reduce the risk from both routes.

Combining the results from previous empirical and modelling research [4, 5, 6, 9, 14] has produced a simple model that aggregates the short-range (i.e., “droplet”) and long-range (i.e., “aerosol”) aerial risks to produce estimates of virion intake.The airpcraft cabin results were generated by visible droplet spray of bactiophage solution that evaporated somewhat in the mock-up cabin environment before measurement at distances of 0.5 to 8 m. While, the generation and transport includes aspects of Routes 1 and 2, the measurements favor aerosol behavior. Aircraft cabin and other environments would be better characterized by more measurements close to infection sources, so that this critical zone could be understood in more detail than what is provided by whole-space decay models.

All of the estmates presented here highlight the importance of masks, which lower the risks to those close to an infected person [1] and at greater distances. Estimates of virion intake depended on the model used (Figures 1 and 2); and, given uncertainties in viral-shedding and in how infection risk scales with exposure duration [8], the continued use of requirements for participating individuals (i.e., masks, vaccination, testing) is justifiable. When the infectivity of nearby occupants is unknown and unchangeableone, such as commercial airplanes or sporting events, perhaps no personal mitigation is available beyond wearing a high-quality mask.

## 5 Conclusion

The importance of droplets and aerosols in SARS-CoV-2 infection have been the focus of debate, and we have provided here some quantification of the relative roles of these routes through literature analysis. Using three published studies, droplets larger than 8 *µm* and smaller than 75 *µm* (medium droplets) and smaller than 8 *µm* (small droplets or aerosols) we have demonstrated that the largest infection risk (as exposure to droplet volume) came from medium droplets, when close to the infectious individual out to approximately 1 m. Larger droplets being most important close to a source comes as no surprise, but verification of this intuition is a step toward focusing public health measures. Farther away, the largest risk was due to aerosols. These trends emerged while summing the contributions of both size-ranges. For one model, medium droplet exposure disappeared at 1.8 m. Policy concerning social distancing for meaningful infection reduction relies on droplet exposure as a function of distance, yet within this construct droplet size determines respiratory deposition. This two-fold distance effect can be used to evaluate additional measures such as plexiglass barriers and masking.

## 6 Weaknesses

Combining the results of multiple studies and simulations as we have done in this paper is far from ideal. The numerical conclusions would be much stronger if all the data were collected in the same way. However, no such study has yet been done, so we were forced to adopt this indirect approach.

## Data Availability

All data produced in the present work are contained in the manuscript

